# A COMPARATIVE STUDY OF DUMPSITES METAL LOADS AND ECOLOGICAL RISK IN SELECTED COMMUNITIES IN KWARA AND EKITI STATES, NIGERIA

**DOI:** 10.1101/2022.12.25.22283930

**Authors:** T.T. Abolayo, H.O. Sawyerr, R.O Yusuf, T.S. Ajai, A.T. Towolawi

## Abstract

This study compared the presence of selected heavy metals in Gbagede dumpsite (GD) and Ilokun dumpsite (ID) dumpsite in Kwara and Ekiti State with the host communities Gbagede community (GC) and Ilokun community (IC). Ten soil samples were collected randomly at 10 cm depth, digested and analyzed for Cadmium (Cd), Copper (Cu), Manganese (Mn), Zink (Zn) and iron (Fe) respectively. The results were compared with control and standard, and subjected to ecological risk indices: Enrichment factor (EF), Contamination factor (CF), and Index of geo-accumulation (Igeo). Metal concentrations were higher (p < 0.05 except Cu) than the control; ranged from 41.2 times (Mn in GD) to 7.08 times (Zn in ID), GD had highest metals except Cu. Metal concentrations at ID > IC (except Cu) and GD> GC. The standard <Cd and Mn,<Zn at GD and ID, and >Cu at GD and GC. The EF was significant (5≤EF<20) for Zn at GD and ID, and for Mn except at GD where there was a very high (20≤EF<40). Mn and Zn indicated high contamination (Cf≥6) at GD and ID while Cd and Fe indicated moderate contamination (1≤Cf<3) except at IC (Cf<1). Moderately polluted (1<Igeo≤2) effects were shown by Fe except at IC and Mn at GD and GC. Conclusively, it was found that the Dumpsites contributed to soil metal loads and ecological risk.

## INTRODUCTION

Increased population and economic development had greatly influenced enormous solid waste generation [1]. The rural-urban drift due to population increase and seeking for a better life has increased the quantity of wastes generated within the urban areas. The receiving counties are thereby facing not only with the challenge of collecting but how and where to dispose of the wastes without further creating environmental hazard [2][3]. Human settlements, small industries and commercial activities contribute greatly to the waste generation dilemma [4][5].The problems are attributed to indiscriminate garbage; roadsides littered with refuse, streams blocked with rubbish and residential areas prone to inappropriately disposed toxic waste from the disposal sites constituting a health hazard [6][7]. Waste is a severe challenge in a developing country like Nigeria due to a lack of awareness of its nature and the skills required to handle it properly [8]. Leachates from refuse dumpsites constitute a source of pollutants not limited to heavy metals on both soil and aquatic environment. Wastes are dumped recklessly with no regards to its environmental implications despite the efforts of waste management authorities in Nigeria. While, municipal wastes are mixed and unsegregated at points of generation, undermining effective management when it comes to treatment and disposal practices. Indiscriminate disposal and sorting of mixed wastes by scavengers have the potential for increased environmental exposure to air, water and soil pollution [9]. The improper waste management also contributes to the contamination of soil while its substantial metal contents are washed away by runoff into water bodies not limited to the streams and rivers thereby making human, animals, plants and the marine environment prone to environmental health risks. Community generated waste is dumped in low lying areas thereby releasing nuisance with resultant threat to natural resources, groundwater and soil [10][11][12].

The health impacts of solid waste generated from households, municipalities, industry, and most importantly, healthcare and hospital facilities on all waste workers cannot be overemphasized. Illegal, uncontrolled and poor disposal of waste threatens the public health of all waste workers, and leads to frequent outbreaks of typhoid, diarrhea, cholera, hepatitis diseases and cancer [13] [12]. After the burning processes that mostly occur on the dumpsite, the heavy metals dissolve in the traversed water and leached into the soil from where they are picked by the components of the biosphere such as growing plants, animals and human downstream. The dissolution sometimes enter the food web generally and food chain specifically. It is known that areas dominated with possible source of metals have the tendencies of higher concentrations of various metals which can lead to environmental pollution [14].

The problem of environmental pollution from toxic metals is a major concern worldwide as they have a deleterious effect on human health and have shown to be hazardous when ingested and or inhaled by humans or via the food chain. Adverse impacts of heavy metals on children had also been proven previously [15][16][17]. Heavy metals may adsorb or adhere to some natural substances, which may increase or decrease their mobility. The transport mechanisms of heavy metals through soil have not been left out by the environmental and soil scientists because of the possible contamination through metal leaching on various spheres of the earth [18] [17].

The impact of solid waste on soil had shown consequently that the presence of and potential exposures of the community to the open dump makes the soil percolation contribute to the tendency of human health being negatively impacted from simple poisoning, cancer, heart diseases and teratogenic abnormalities [19]. Soil contamination through indiscriminate waste discharges is undoubtedly a challenge worldwide with carriage of different metals transferred to and up taken by biosphere in different ways [20].The metal pollution of the environment, even at low levels, results in long-term cumulative health effects and it is among the leading health concerns all across the world [21][17]. Thus, the current research aimed at comparing soil heavy metal contents around two waste dumpsites and their respective community; Gbagede in Ifelodun Local Government Area of Kwara State and Ilokun in Irepodun Local Government Area of Ekiti State. The former dumpsite is located within 0-50m while to residential area while the latter is located within 50-100m to highly density residential areas as development is fast approaching the dumpsite. However, an important aspect of this work was comparison with the world-soil standards and adoption of ecological health risk indices using background values of heavy metals.

## MATERIALS AND METHODS

### Study area

This study was carried out on two dumpsites and the respective adjoining community; Gbagede located in Ifelodun Local Government Area (LGA) of Kwara State in North-central Nigeria and Ilokun located in Irepodun LGA of Ekiti State, Southwest in Nigeria. The dumpsites were located within 0 - 50m in Gbagede and 50 – 100m in Ilokun to the high density residential area as development is fast approaching the dumpsites. However, the necessary safety distance between human settlements and waste landfills must be at least 1,000 m and that between landfills and forests, protected areas and watercourses, must be at least 500 m had been violated here. Field survey was undertaken on 30th October, 2020 to collect point data (coordinates) of the study area and map the dump site locations. The point data was collected using Garmin GPS. The geo-coordinates were initially collected in Degrees, Minutes and Seconds (DMS) and converted subsequently into Degrees Decimals (DD) using Geo-calculator.

To delineate and identify the major land use in the study area, satellite images of the study area were obtained by downloading a base map using the new feature of Arc GIS 10.4 version. Figure 1 shows maps of the two LGAs with the red mark indicating the dumpsites with their respective adjourning community. The dumpsites studied contain mixture of both organic and inorganic waste materials such as food wastes, papers, metals, tins, glass, ceramics, battery wastes, textile materials, plastics, ash, fine dust, rubber, wood wastes, sewage and other miscellaneous materials.

**Figure 1:**
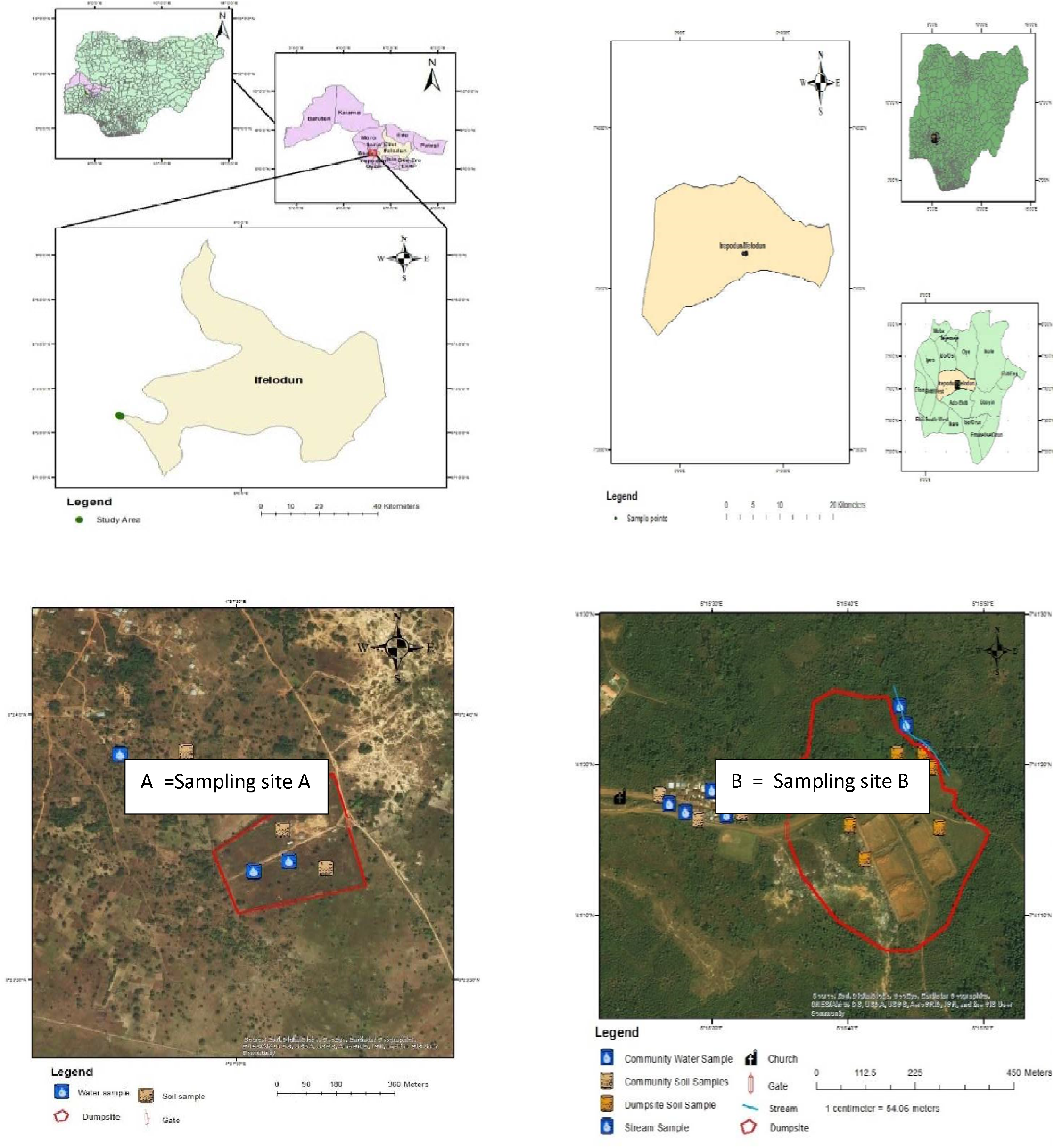
Map of the study areas showing sites A (Kwara State) and B (Ekiti State).

### Soil sampling

Surface soils are the first locus of input of metals where they tend to accumulate on a relatively long term basis, 10 surface soil samples were collected randomly to a depth of 0-20cm from each of the two designated dumpsites and bulked respectively for a representative sample. For comparison of the results, soil samples of uncontaminated area to a depth of 0-20cm over 100 m away to Gbagede (Kwara State) and 200m away to Ilokun (Ekiti State) from the contaminated (open dump) site were also collected randomly and bulked. This is because of their respective distance from the dumpsites to the adjourning community. The soil samples were placed in labeled polythene bags and transported to the laboratory. All the soil samples were air-dried subsequently to constant weight [22]. They were homogenized, made lump free by gently crushing repeatedly using an acid pre-washed mortar and pestle, and passed through a 1.5mm plastic sieve prior to digestion [23].

### Soil heavy metal contents analysis

The soil cadmium (Cd), copper (Cu), lead (Pb), zinc (Zn) and Iron (Fe) contents were analyzed using atomic absorption spectroscopy after nitric acid (HNO3)^-^ and hydrogen peroxide (H_2_O_2_) digestion [24]. The soil sample was thoroughly mixed to achieve homogeneity and sieved using a USS #10 sieve. For each digestion procedure, a 10ml of 1:1 HNO_3_ was added to the 1 g of the sample and covered with a watch glass. The sample was heated to 95 ± 5°C and refluxed for 10 to 15min without boiling. The sample was allowed to cool, 5mL of concentrated HNO_3_ was added, with the cover replaced, and refluxed for 30 min. Brown fumes evolved to indicate oxidation of the sample by HNO_3_. The step was repeated by (addition of 5mL of conc. HNO_3_) two to four times until no brown fumes gave off as an indication of complete reaction with HNO_3_. The solution evaporated to approximately 5mL by heating at 95 ± 5°C without boiling for 2 h. After the aforementioned step completed and the sample cooled, 2mL of water and 3mL of 30% H_2_O_2_ were added. The vessel was covered and the peroxide reaction initiated, 1mL of 30% H_2_O_2_ was added continuously with warming until the effervescence was minimal or the general sample appearance was unchanged. The sample was again covered with the ribbed watch glass device while the acid-peroxide digestate continued until the volume reduced to approximately 5mL while being heated at 95 ± 5°C without boiling for 2h. The digestate was filtered through Whatman No. 41 filter paper after cooling; the filtrate was collected in a 100mL volumetric flask and made to volume for analysis. The analytical phases involved analysis of the soil heavy metal concentrations around the dumpsites and their communities using Atomic Absorption Spectrophotometer (AAS).

### Statistical Analysis

The obtained data were analyzed using IBM SPSS Statistics 22 for descriptive (mean and standard deviation, tabulation and chart) and inferential (DMRT) statistics for the level of significance at p < 0.05.

### Quantification of Soil Pollution using the Ecological Risk Indices

Results of the analyzed data were compared with that of samples from the control points (as the background/ uncontaminated values) to have an idea about the soil heavy metal contents (as the contamination levels) around the dumpsites. The background value of an element is the concentration of that element obtained from a control site investigated and proven to have not been disturbed by the anthropogenic (human) activities under consideration [25]. Various quantitative indices have been employed to assess the impact of anthropogenic (human) activities on the concentration of toxic/ trace metals in the soil for ecological risk assessment, and among such indices are the three (enrichment factor, contamination factor and index of geo-accumulation) used in this study. They are as described below

### Enrichment factor (EF)

An index called Enrichment factor (Ef) was initially developed to speculate origin of the elements in the atmosphere or precipitation [26] but was progressively extended to the study of soils, lake sediments, peat, tailings and other environmental materials [27]. In this study, Ef was used to assess level of contamination for the possible dumpsite impacts on the study soil samples. The Ef was calculated according to the equation generalized by [26] as:

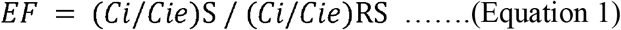

Where *Ci* is the content of element *i* in the sample of interest (S) or the selected reference sample (RS), and *Cie* is content of immobile element in the S or RS. So (*Ci/Cie*)S is the heavy metal to immobile element ratio in the S, and (*Ci/Cie*)RS is the heavy metal to immobile element ratio in the RS. The RS is usually an average crust or a local background sample. The immobile element is often taken to be Al, Li, Sc, Zr, or Ti [26]. Sometimes Fe or Mn has been used. The EF values to be interpreted as EF <2 (deficiency to minimal enrichment), 2<EF<5 (moderate enrichment), 5<EF<20 (significant enrichment), 20<EF<40 (very high enrichment) and EF>40 (extremely high enrichment) [27].

### Contamination factor (Cf)

An assessment index is generally applied to measure environmental quality of soil and one simple and well-known single element pollution index is the Contamination factor (Cf). The Cf is used to describe contamination of a given toxic substance in either an aquatic or a terrestrial environment. In calculating Cf, the equation suggested by [28] was used as follows:

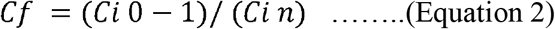

Where *Ci*_*0-1*_ is the mean content of the determined metal and *Ci*_*n*_ is the pre-industrial concentration of the metal, which is the concentration of the control (uncontaminated) sample suggested by [29], in this study. The following gradations were proposed: Cf<1 low contamination; 1≤Cf<3 moderate contamination; 3≤Cf<6 considerable contamination; Cf<6 high contamination.

### Index of geo-accumulation (Igeo)

An index of geo-accumulation (Igeo) was originally defined by [30] in order to determine and define metal contamination in sediments by comparing current concentrations with pre-industrial levels. Index of geo-accumulation can be calculated by the following equation:

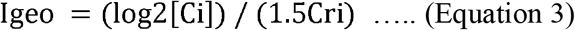

Where Ci is the measured concentration of the individual determined metal i and Cri is the geochemical background concentration or reference value of the metal i, which had been taken to be the concentration of the determined metal in the control sample. Factor 1.5 is used because of possible variations in background values for a given metal in the environment and or very small anthropogenic influences. Muller classified Igeo index as unpolluted (Igeo≤0), unpolluted to moderately polluted (0<Igeo≤1), moderately polluted (1<Igeo≤2), moderately to heavily polluted (2<Igeo≤3), heavily polluted (3<Igeo≤4), heavily to extremely polluted (4<Igeo≤5), and extremely polluted (Igeo>5).

## RESULTS AND DISCUSSION

Summary of the soil heavy metal contents across the two dumpsites Gbagede in Kwara (GD) and Ilokun in Ekiti States (ID) and their adjoining communities are as presented in Figures 2 and 3 with the values of soil heavy metal contents compared with both the control soil samples’ values and the world-soil average given by [31]. Table 1 computed Ecological Risk indices: Contamination factor (Cf), Enrichment factor (Ef) and Index of geoaccumulation (Igeo). With the exception of Fe, concentrations of the metals in dumpsites are from 41.2 times (for Mn in GD) to 7.08 times (for Zn in ID) higher than that of the control samples. Concentrations of the metals (except Cu) were highest in GD, higher across the study than control sites. Concentrations of Cd, Mn, and Fe were higher at GC while Cu was highest at the IC. There was an observation that soil metal contents of GD> GC, ID > IC (except Cu), while Cd and Mn > the compared standard. The Zn soil content at GD and ID > while Cu soil content at GD and GC < the compared standard, all the soil metal contents varied significantly (p < 0.05) except Cu (p > 0.067).

**TABLE 1.**
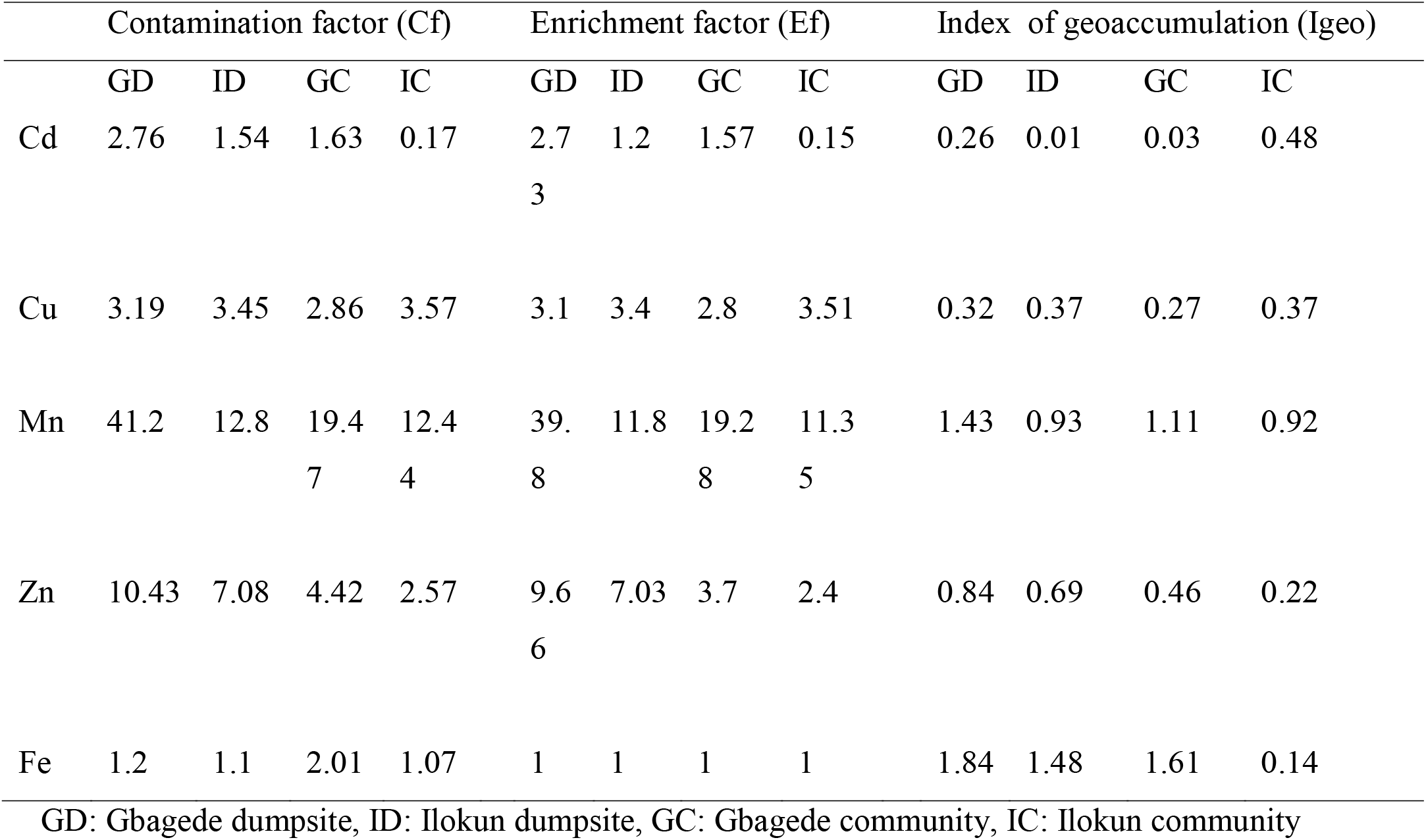
Ecological risk indices

**Figure 2:**
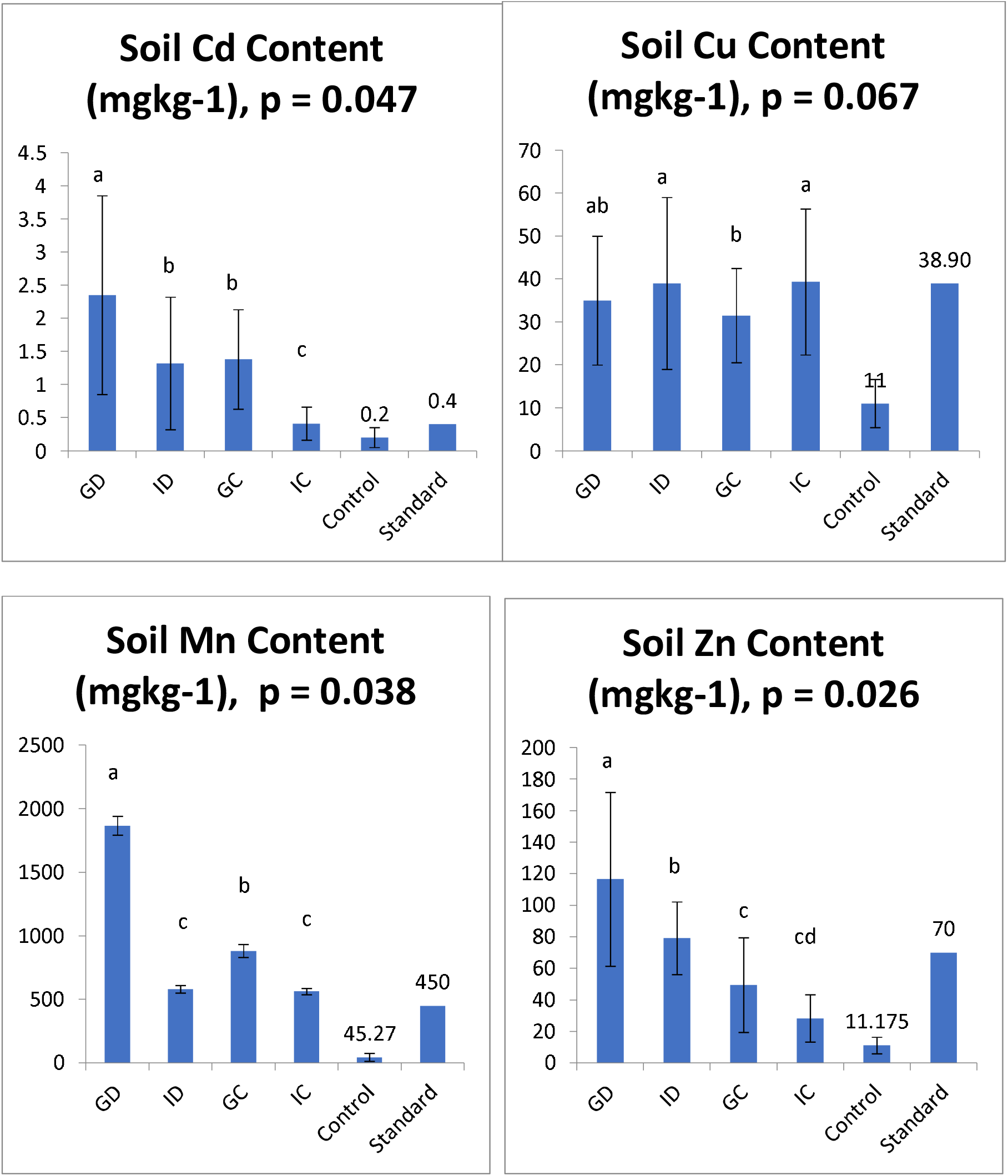
Concentrations (mgkg^-1^) of heavy metals in the soil samples. Gbagede dumpsite (GD), Ilokun dumpsite (ID), Gbagede community (GC), Ilokun community (IC).

**Figure 3:**
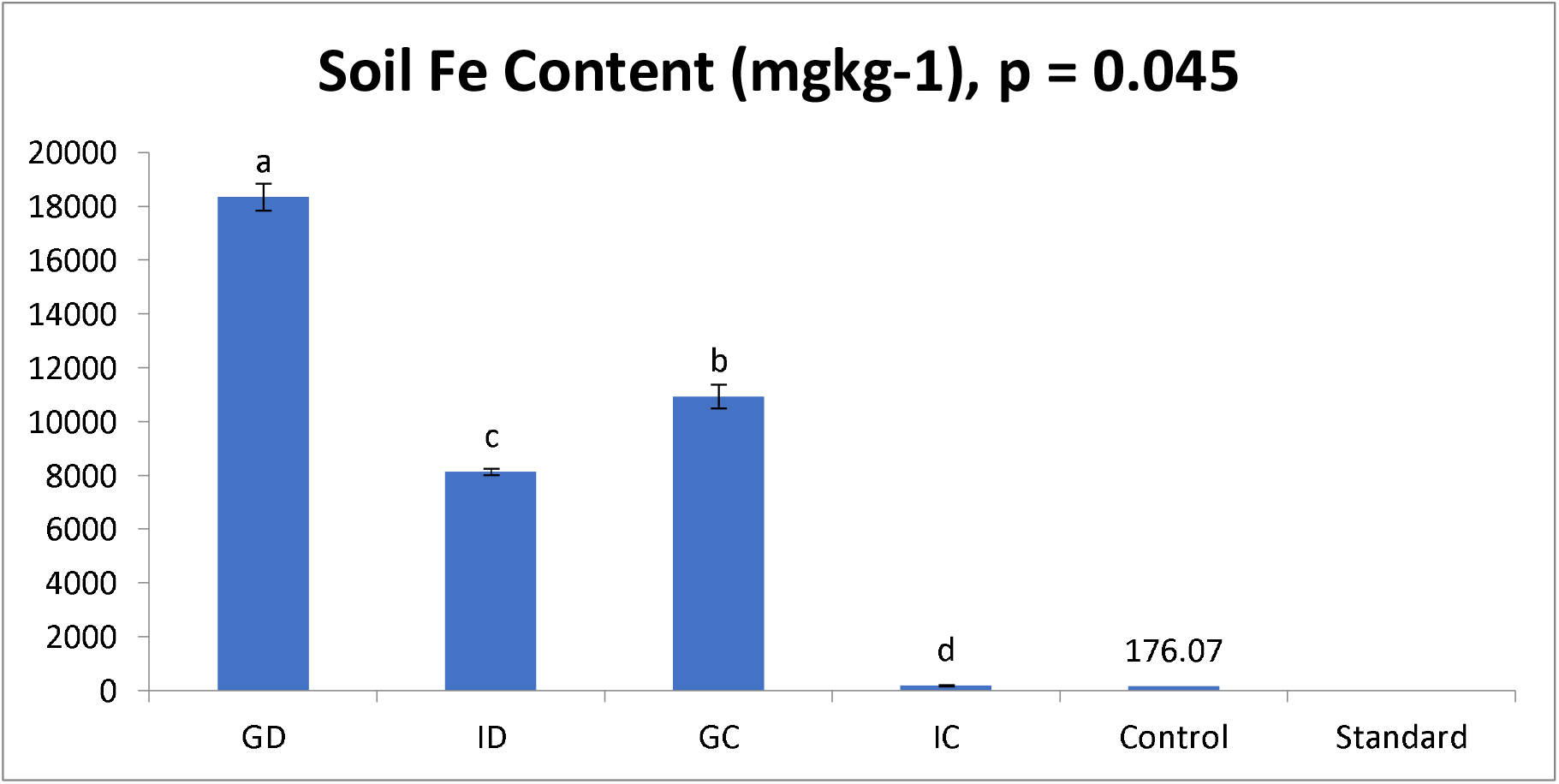
Concentrations (mgkg^-1^) of heavy metals in the soil samples.Gbagede dumpsite (GD), Ilokun dumpsite (ID), Gbagede community (GC), Ilokun community (IC).

### Soil heavy metal contents across the study areas

A comparison was made for the levels of heavy metals in the (Gbagede, GD and Ilokun, ID) dumpsites and Gbagede community, GC and Ilokun community IC with the world-soil average standard given by [31]. Many studies have shown that urban soil receive load of contaminants due to the anthropogenic (human) activities in the urban settlements more than the sub-urban or rural areas nearby [32]. This is corroborated largely by this study shown in the concentrations of the investigated (GD, ID, GC and IC) metals in the dumpsites and control soils. The mean levels of Cd exceeded the standard of approximately 0.40 mgkg^-1^ given by [31] with an indication of anthropogenic sources on dumpsites with possibility of affecting the neighboring community soil, it has been reported that inputs of Cd into soils may be of different origins such as agricultural amendment, sludge from decomposed wastes and atmospheric deposition [8].Cadmium has a wide range of uses in the industry that deal in paints, pigments, electroplating and plastic stabilizer [29] [8]. Many other anthropogenic activities that can increase soil Cd above the background levels are not limited to the burning of fossil fuel and tires, use of lubricating oils, vehicle wheels, and deposition of solid wastes from industries and home, sewage sludge, wastewater irrigation and phosphate fertilizer application [33].

All these end-up on the site to make the dumpsite soil and the nearby community contaminated with high Cd compare with standard. Cadmium (Cd) is an element of great concern from toxicity point of view with its exposure causing both chronic and acute health effects in living organisms, having terrestrial abundance of 0.1 - 0.2 mg/kg on average, its compounds are carcinogenic to humans and are classified as Group 1 by International Agency for Research on Cancer as the element and its compounds are cancerous to the lung, kidney and of the prostate [34]. Its intoxication leads susceptibly to pulmonary damages, kidney damage, skeletal damage, and itai-itai diseases [35] [36].

Natural and anthropogenic sources (mine/ smelter wastes, phosphate fertilizers, or sewage sludge, and municipal waste landfills) contribute note worthily to the levels of Cd in the soil and sediments [34]. Observation had it that there could be a substantial variation in Cd levels ranging from 0.02 and 184 mg/kg from normal soil to the contaminated soil with diverse activities [37] [38]. The study of [39] determined highest concentrations of Cd in the soil to be 6.74mg/kg, suggested its high portion entering higher food chain and reflected by the forage Cd accumulation in the range of 1.14 to 4.20mg/kg.

The observed mean levels of Cu for ID and IC were higher than in the control sample and standard reported by [31]. However, Cu concentrations in GD and GC were within the standard. The element (Cu) acts as redundant in the enzymes superoxide dismutase, cytochrome oxidase, lysyl oxidase, dopamine hydroxylase, and several other oxidases that reduce molecular oxygen. It is transported in the organism by the protein ceruloplasmin and recommended at 0.9mg/day in dietary allowance for adults [40]. The USA has its median dietary intake of Cu at 1.0 to 1.6 mg/day approximately with its tolerable upper intake level at 10 mg/ day for adults [41]. This indicated that the soil samples under this current study had a very high concentration of Copper (Cu) Manganese (Mn) are among the most abundant elements in the earth crusts and distributed widely in soils, sediments, and rocks [42]. Although the observed values were all higher than the standard, studies carried out by [31] and [43] reported Mn values higher than the values reported in this current study. Comparison with the control samples indicated that the current obtained Mn concentrations were 14 times higher than the control soil contents.

The Zn determined values were within the values given 10 - 300 mgkg^-1^ by [44], but higher in two dumpsites (GD and ID) than the standard 70.00 mgkg^-1^ and soil contents of the communities that host the dumpsites. Environmental contamination of Zn is mainly related to anthropogenic input. As corroborated that the anthropogenic sources of Zn are related to industries dealing in liquid manure, composted materials and agrochemicals such as fertilizers and pesticides in agriculture [31]. The chemical (Zn) may be derived from mechanical abrasion of vehicles as they are used in the production of brass alloy itself and come from brake linings, oil leak sumps and cylinder head gaskets [45]. Zinc is essential micronutrient and catalyzes enzyme activity, contributes to protein structure, and regulates gene expression [41]. Although consequent Zn deficiency was recognized for many years, it can be toxic when exposures exceed physiological needs [46] with acute gastrointestinal effects and headaches, impaired immune function, changes in lipo protein and cholesterol levels, reduced copper status, and zinc iron interactions [47]. The RDA of Zn for adults is between 8 and 11mg/day(female-male), whereas the tolerable upper intake level is 40mg/day for adults, a value based on reduction in erythrocyte copper-zinc superoxide dismutase activity [47].

The mean metal concentrations of Fe in the current study soil samples were all higher than the range of values reported previously 1100 and 10,920 mgkg^-1^respectively [48] [49]. The high concentrations of Fe determined in this current study hardly seemed to pose threat on the dumpsites surroundings considering the report of past study that Fe had effect on the plant growth as easily soluble and exchangeable fractions of Fe were very low in comparison with the total Fe content in soil [31]. Having a range of Fe content in soil from different regions was between <1 and 196mg/ kg. A remarkably high mean value of 25080 to 26960mg/kg Fe had been reported in contaminated soil [50]. Such values were higher than the values determined in this current study. Iron is important in human body metabolism acting as a catalyst and occurring greater than any other trace element and functioning as a component of a number of proteins such as enzymes and hemoglobin. However, the human RDA is 8mg/ day and tolerable upper intake for adults is 45mg/ day Fe, which is based on gastrointestinal distress as an adverse effect [41].

The high soil metal contents determined in this study showed that soil of the considered communities are currently under encroachment of heavy soil metals pollution and serious threats of metal epidemic. A direct relationship existing between anthropogenic activities, waste load, and high metal pollutant of a place as shown by this study meant that a greater number of Nigerian urban cities are susceptible to the threat of metal pollution, given the mountainous heaps of refuse and other complex wastes commonly sighted in most cities. Environmentally, metal poisoning is a silent epidemic, so findings of this study suggest that every resident of GC and IC might be at risk of metal epidemic which is possible to have been ravaging lives silently and already.

### Ecological risk assessment around study areas

The concentrations of the control (uncontaminated) samples were taken as the reference concentrations while Fe was taken as the immobile element as proposed by [30]. Iron (Fe) had been an acceptable immobile (normalization) element used in the *Enrichment factor (Ef)* calculation since they considered Fe distribution to be unrelated to other heavy metals [53]. Thus to determine the relative degree of metal contamination, comparisons were made to background concentrations using Fe as the immobile element following the assumption that its content in the crust has hardly been disturbed by anthropogenic activity, thereby being chosen as the element of normalization because natural sources (98%) dominate vastly its input [51]. The process of standardization helps in evaluating the anthropogenic component over and above the natural component. When juxtaposed with categories of enrichment factor invented by [52], there occurs a somewhat different conclusion. For the GD, there was a minimal enrichment (EF<2) for Fe and Cd except at GD, moderate enrichment (2≤EF<5) for Cu, significant enrichment (5≤EF<20) for Zn at GD and ID, and for Mn except at GD where there was an indication of a very high enrichment (20 ≤ EF < 40). The observed Ef of 1.00 for Fe was expected because it served as the normalization element in the enrichment factor calculation.

The *Contamination factors* (Cf) can be used to differentiate between the metals originating from anthropogenic activities and natural processes to assess the degree of anthropogenic influence [54]. The Cf of the heavy metals at the two dumpsites (GD and ID) and their adjourning communities (GC and IC) as compared with the various categories of contamination factors showed that there was high contamination (Cf ≥6) of Mn across the four sites but of Zn in GD and ID; considerable contamination (3≤Cf<6) of Cu except at GC, where there was an indication for Zn; both Cd and Fe were determined to indicate moderate contamination (1≤Cf<3) across the four sites. High Cf suggests a strong anthropogenic influence.

The degree of metal pollution was assessed with respect to the seven contamination classes known as *Index of geoaccumulation (Igeo)* [26][55]. Results of the current study showed that moderately polluted (1<Igeo≤2) effects were quantified for Fe except at IC while Mn at GD and GC. However, both dumpsites (GD and ID) and their respective community (GC and IC) soil samples had been unpolluted (Igeo≤0) by Cd, Cu and Zn.

## Data Availability

All data produced in the present work are available upon reasonable requests to the authors

## CONCLUSION AND RECOMMENDATIONS

Comparison between Gbagede and Ilokun dumpsites indicated different soil metals (Cd, Mn, Fe and Zn) contents except in Cu which had no significant (p = 0.062, > 0.05) difference. It was indicated that significant differences of other metals occurred across the four study sites and might be due to composition and sources of the dumped wastes. The high soil metal (Zn, Cd, Cu, Mn, Fe) contents in and around the dumpsites could be traced to the poor management and indiscriminate open dumping of solid wastes. High concentrations of the metals overtime would make the environment susceptible to metal pollution, soil metal overload, and harmful effects from metal toxicity which in turn affect plant, animals and human.

The ecological risk indices showed high contamination (Cf≥6) of Mn and Zn; considerable contamination (3≤Cf<6) of Cu in both dumpsites and Ilokun community; but moderate contamination (1≤Cf<3) of Cd and Fe. At the Gbagede dumpsite (Kwara State), there was a very high enrichments (20≤EF<40) for Mn; significant enrichment (5≤EF<20) for Zn; moderate enrichment (2≤EF<5) for Cu and Cd; minimal enrichment (EF< 2) for Fe. At the Ilokun dumpsite (Ekiti State), there was a significant enrichment (5≤EF<20) for Zn and Mn; moderate enrichment (2≤EF<5) for Cu; but minimal enrichment (EF<2) for Fe and Cd. For the Gbagede community, there was a significant enrichment (5≤EF<20) for Mn; moderate enrichment (2≤EF<5) for Cu and Zn; but minimal enrichment (EF<2) for Cd and Fe. For the Ilokun community, there was a significant enrichment (5≤EF<20) for Mn; moderate enrichment (2≤EF<5) for Cu and Zn; but minimal enrichment (EF<2) for Fe and Cd.

